# Reciprocal longitudinal associations between symptoms of eating disorders, self-harm and suicidal ideation

**DOI:** 10.1101/2025.03.19.25324248

**Authors:** Agnieszka Musial, Una Foye, Saakshi Kakar, Tom Jewell, Janet Treasure, Gursharan Kalsi, Iona Smith, Laura Meldrum, Shannon Bristow, Ian Marsh, Chelsea Mika Malouf, Jahnavi Arora, Helena Davies, Ellen J. Thompson, Rina Dutta, Ulrike Schmidt, Gerome Breen, Moritz Herle

## Abstract

**Background:** Eating disorders are severe psychiatric conditions associated with high mortality rates, particularly among young people. These disorders often co-occur with self-harm and suicidal ideation, yet the temporal dynamics between these variables remain poorly understood.

**Aims:** This study aims to elucidate the longitudinal associations between eating disorder symptoms, self-harm, and suicidal ideation using structural equation modelling.

**Method:** Repeated measures of these phenotypes were used to construct a hypothetical model that includes cross-path analyses within and between the variables in two cohorts: the Twins Early Development Study (TEDS; ages 16, 21 and 26; *N*=5,196), representing a general population sample, and the Covid-19 Psychiatry and Neurological Genetics study (COPING; data collected between June 2020 and July 2021; N=490), which focused on individuals with a history of anxiety or depression. In the TEDS cohort, symptoms of eating disorders, self-harm, and suicidal ideation showed limited continuity across adolescence and young adulthood, with peak symptom severity at age 21.

**Results:** Cross-domain associations revealed that both self-harm and suicidal ideation at age 21 were more strongly associated with eating disorders at 26 than the reverse. In contrast, the COPING cohort exhibited more stability in symptoms over time but showed minimal cross-domain effects.

**Conclusions:** The effects of self-harm and suicidal ideation on eating disorders in early adulthood are stronger than the influence of disordered eating on suicidality.

## Introduction

Eating disorders, including anorexia nervosa (AN), bulimia nervosa (BN), and binge eating disorder (BED), are associated with some of the highest mortality rates among psychiatric illnesses, resulting in over 3.3 million premature deaths annually ^1–4^. A significant number of eating disorder-related deaths are due to suicide ^3,5–7^, particularly among AN patients (one in five deaths) ^8^. The prevalence of suicidal ideation in individuals with AN and BN had been estimated as ranging between 50% and 60% and between 17% and 25% for suicide attempts ^7^. Research presents mixed findings on the mechanisms linking eating disorders and suicidality. Some studies suggest symptoms of eating disorders lead to suicidal thoughts and behaviours, while others propose that suicidality might precede or even contribute to the development of eating disorders and that both conditions may share biological and psychological risk factors ^9^. Limited longitudinal research has examined how eating disorder symptoms predict suicide outcomes, though a meta-analysis of 14 studies found eating disorders significantly predict suicide attempts, but not death ^10^. Some studies also suggest that symptoms of eating disorders can arise following suicidal thoughts and attempts, highlighting a potentially bidirectional relationship ^11,12^. A common precursor to suicide in the general population is self-harm, with about 60% of individuals who complete suicide having a history of self-harm, mostly within a year before the suicide attempt ^13–15^. According to a meta-analysis of 29 studies, individuals with eating disorders are prone to engage in self-harm, with 22% of AN and 33% of BN patients having reported lifetime self-harm ^16^. A more recent systematic review estimates the prevalence of non-suicidal self-injury in eating disorder patients as even higher—40% ^7^. Due to the widely demonstrated co-occurrence between eating disorders, self-injury and suicidal ideation and attempts, The National Institute for Health and Care Excellence (NICE) has emphasised the need for research focusing on psychiatric conditions co-occurring with eating disorders, with a particular focus on self-harm (NICE, 2017). This urgency is amplified by the co-occurrence between eating disorders and self-harm being ranked among the top 10 research priorities in a survey of patients, carers, and clinicians ^18^ and by the COVID-19 pandemic and associated lockdowns, which have led to an increase in new eating disorder diagnoses ^19,20^. Currently, no comprehensive theoretical framework exists to explain why individuals with a history of eating disorders face an elevated suicide risk. A relevant model in this area is the interpersonal-psychological theory of suicide, which suggests that suicide risk arises when three conditions are present: a sense of social isolation or lack of belonging, feelings of being a burden to others, and an acquired capability for suicide ^21^. While this theory has been influential, evidence on its applicability to eating disorders remains sparse. A recent review identified 10 small, mostly cross-sectional studies that examined this model in the context of eating disorders, highlighting a pressing need for large-scale, longitudinal research with extended follow-up and repeated measures to more effectively assess suicide risk in eating disorder patients ^22^. The aim of the present project is to investigate the longitudinal associations between symptoms of eating disorders and subsequent self-harm and suicidal ideation, utilising structural equation modelling framework in two large UK-based cohorts, Twins Early Development Study (TEDS) and the COVID-19 Psychiatry and Neurological Genetics (COPING) study. These cohorts differ substantially in their profiles, allowing us to explore the temporal dynamics between symptoms of eating disorders, self-harm and suicidal ideation between a developmental, population-based sample (TEDS) and cross-sectional clinical sample (COPING). The present study seeks to provide a comprehensive understanding of how eating disorder symptoms contribute to the development of suicidal ideation.

## Method

### Sample

The current study used data from two United Kingdom based cohorts, the TEDS ^23^, which is a population-based twin sample and the COPING study ^24^, which is a clinical cohort, recruiting predominantly participants with a lifetime history of anxiety and mood disorders.

### Twins Early Development Study (TEDS)

The TEDS sample consists of more than 10,000 pairs of twins born in England and Wales between 1994 and 1996. The TEDS twins have been assessed a dozen times from infancy through early adulthood on a variety of demographic, behavioural, cognitive and mental health measures. The sample of TEDS twins is representative of the UK population in terms of ethnicity and socioeconomic status (SES); for details of representativeness and attrition, see ^25,26^. In the present study we investigated the longitudinal relationships between symptoms of eating disorders measures at ages 16, 21 and 26 and measures of self-harm and suicidal ideation collected at ages 21 and 26. The mean age of the sample was 16.3 years (*SD*=0.69) at age 16 data collection, 22.3 years (*SD*=0.91) at age 21 data collection and 26.4 years (*SD*=0.92) at age 26 data collection. The sample selected for analyses included 5,196 individuals, out of whom females comprised 67% (*N*=3,468) and 95% (*N*=4,931) of participants reported European ethnic origin. Within the sample, 33% (N=1718) of individuals reported experiencing lifetime depression and 20% (N=1026) reported experiencing lifetime anxiety. Out of the total 5,196 twins, 3% (N=172) of individuals were diagnosed with AN, 2% (N=82) individuals were diagnosed with BN and 1% (N=48) of individuals were diagnosed with BED.

### COPING

The sample included participants from the National Institute for Health and Care Research (NIHR) BioResource who joined the COPING study ^24^. Alongside COVID-related measures, the COPING study incorporated questionnaires from the Genetic Links to Anxiety and Depression (GLAD) Study and the Eating Disorders Genetics Initiative UK (EDGI UK) ^27,28^. For further information on the sub-cohorts, recruitment and exclusion criteria, please refer to ^24,29^. For the present analysis, we used data on symptoms of eating disorders, self-harm and suicidal ideation collected at first follow-up, that is in early June 2020 (referred to as baseline henceforth), eighth follow-up in September/October 2020 and twentieth follow-up in June/July 2021. We additionally included data related to psychosocial aspects of participants’ lives, collected at the third follow-up in late June 2020. Our selected sample resulted in a total of 490 individuals with complete data for at least one of the timepoints for symptoms of eating disorders, self-harm and suicidal ideation. The mean age of the sample was 47.7 years (*SD*=15.9). Females comprised 53% (*N*=261) of the sample and 81% (*N*=397) of participants reported European ethnic origin. Within the selected sample, 42% (N=204) of individuals self-reported lifetime history of major depression and 33% (N=163) of individuals reported lifetime history of anxiety disorder. Regarding lifetime eating disorder diagnoses, 5% (N=24) of individuals reported being diagnosed with AN, 2% (N=9) of individuals reported a diagnosis of BN and 1% (N=7) of individuals reported a diagnosis of BED. In addition, four individuals reported being diagnosed with purging disorder, seven with avoidant/restrictive food intake disorder and two with other feeding or eating disorder.

### Ethical approval

TEDS has ethical approval from Kings College London Research Ethics Committee (References: PNM/09/10–104 and HR/DP-20/21–22060). Consent was obtained before data collection at every wave.

The London—Fulham Research Ethics Committee approved the GLAD Study on 21st August 2018 (REC reference: 18/LO/1218) and EDGI UK on 29th July 2019 (REC reference: 19/LO/1254). The NIHR BioResource has been approved as a Research Tissue Bank by the East of England— Cambridge Central Committee (REC reference: 17/EE/0025). The COVID-19 Psychiatry and Neurological Genetics study was approved by the South West—Central Bristol Research Ethics Committee on 27th April 2020 (REC reference: 20/SW/0078). The RAMP Study was approved by the Psychiatry, Nursing, and Midwifery Research Ethics Committee at King’s College London on 27th March 2020 (HR-19/20–18157).

### Measures

#### Symptoms of eating disorders

In the TEDS sample, different measures of seating disorders symptoms were administered at each age. At age 16, the twins responded to four items from the Eating Disorder Diagnostic Scale (EDDS) ^30^, asking about body perception during the previous six months. At age 21, the 12-item Eating Disorder Inventory-2 ^31^ was administered, encompassing items related to the restricting, bingeing/purging and body image. At age 26, eating disorders symptoms were measured using a screener of AN, BN and BED symptoms, mapping onto DSM-5 criteria ^32^. All items are listed in Supplementary Table 1. In the COPING sample, symptoms of eating disorders were assessed using a 13-item Eating Disorder Examination Questionnaire (EDE-Q) measure, which is a brief measure of eating disorder symptom severity ^33^. In addition to the total EDE-Q score, we also created symptom scores indexing restricting, purging and bingeing behaviours. These symptoms scores were created as latent variables using structural equation modelling (SEM) in *lavaan* as a data reduction technique ^34^. For a list of items and details on symptom score items please refer to Supplementary Table 2.

#### Self-harm and suicidal ideation

In the TEDS cohort, at age 21, self-harm behaviours were measured using 10 items adapted from the Child & Adolescent Self-harm in Europe (CASE) Study ^35^ and ^36^. Suicidal ideation was measured using 3 items adapted from the same surveys ^35,36^. At age 26, self-harm and suicidal ideation were assessed using the following two Likert-scale scored items, *In the past year, have you ever hurt or harmed yourself on purpose in any way (e*.*g*., *by taking an overdose of pills, or by cutting yourself)?* and *In the past year, have you ever thought about killing yourself, even if you would not really do it?* ^35,36^. Self-harm and suicidality data was not collected at age 16 in TEDS. In the COPING sample, self-harm and suicidal ideation were measured using the Thoughts and Feelings questionnaire (TAF) ^37^.

The following two items were used to assess symptoms of self-harm: from the: *Have you contemplated harming yourself?* and *Before the pandemic, had you deliberately harmed yourself, whether or not you meant to end your life?*. Suicidal ideation was assessed by the following item: *Many people have thoughts that life is not worth living. Have you felt that way?*. The remaining items temporally related to the COVID-19 pandemic were discarded.

#### Mental health diagnoses

In TEDS, data on lifetime depression and anxiety were collected using the Composite International Diagnostic Interview for Depression (CIDID) and Anxiety (CIDIA) ^38^. Eating disorder diagnoses were assessed using a bespoke measure developed by the Psychiatric Genomics Consortium (PGC) for the UK Biobank study. In COPING, diagnoses of eating disorders, major depressive disorder and generalized anxiety disorder were evaluated based on the Mental Health Diagnosis questionnaire (MHD), adapted from the UK Biobank Questionnaire ^39^. This questionnaire was integrated into the baseline COVID assessment for the remaining COPING participants.

#### Analyses

Analyses for this project were preregistered with the Open Science Framework (OSF) (https://osf.io/xy273/). The hypotheses are listed in Supplementary Note 1.

#### Longitudinal models

A structural equation modelling (SEM) framework was employed to explore the longitudinal associations between symptoms of eating disorders, self-harm, and suicidal ideation. Repeated measures of these variables were used to construct a hypothetical model that included cross-path analyses within and between the variables, as illustrated on Figure 1. In TEDS, we investigated the temporal relationships between symptoms of any eating disorder measured at ages 16, 21 and 26 and symptoms of self-harm and suicidal ideation measured at ages 21 and 26, using all available data from the twins and accounting for family clustering. An equivalent model was estimated in the cross-sectional COPING cohort, where symptoms of specific eating disorders, including AN, BN and BED were measured at baseline, followed by two follow-up assessments of any eating disorder administered five and eight months apart. Data on self-harm and suicidal ideation were collected using the same instruments at baseline, as well as the follow-ups. Covariates included age at data collection, sex and socioeconomic status (SES) in TEDS and age at baseline and sex in COPING. In both cohorts, we restricted the sample to those individuals who had complete data for at least one timepoint of each variable and subsequently used the full information maximum likelihood (FIML) implemented in *lavaan* to account for data missingness. We repeated the analyses using a subset of individuals diagnosed with AN, BN or BED. We have additionally performed the mediation analyses between symptoms of eating disorders at baseline and suicidal ideation at the final timepoint employing candidate mediators that included mid-point measures of psychosocial variables related to interpersonal relationships, health behaviours and life events. For details and results of the mediation analyses please refer to Supplementary Note 2, Supplementary Table 5 and Supplementary Figures 3 and 4.

**Figure 1.**
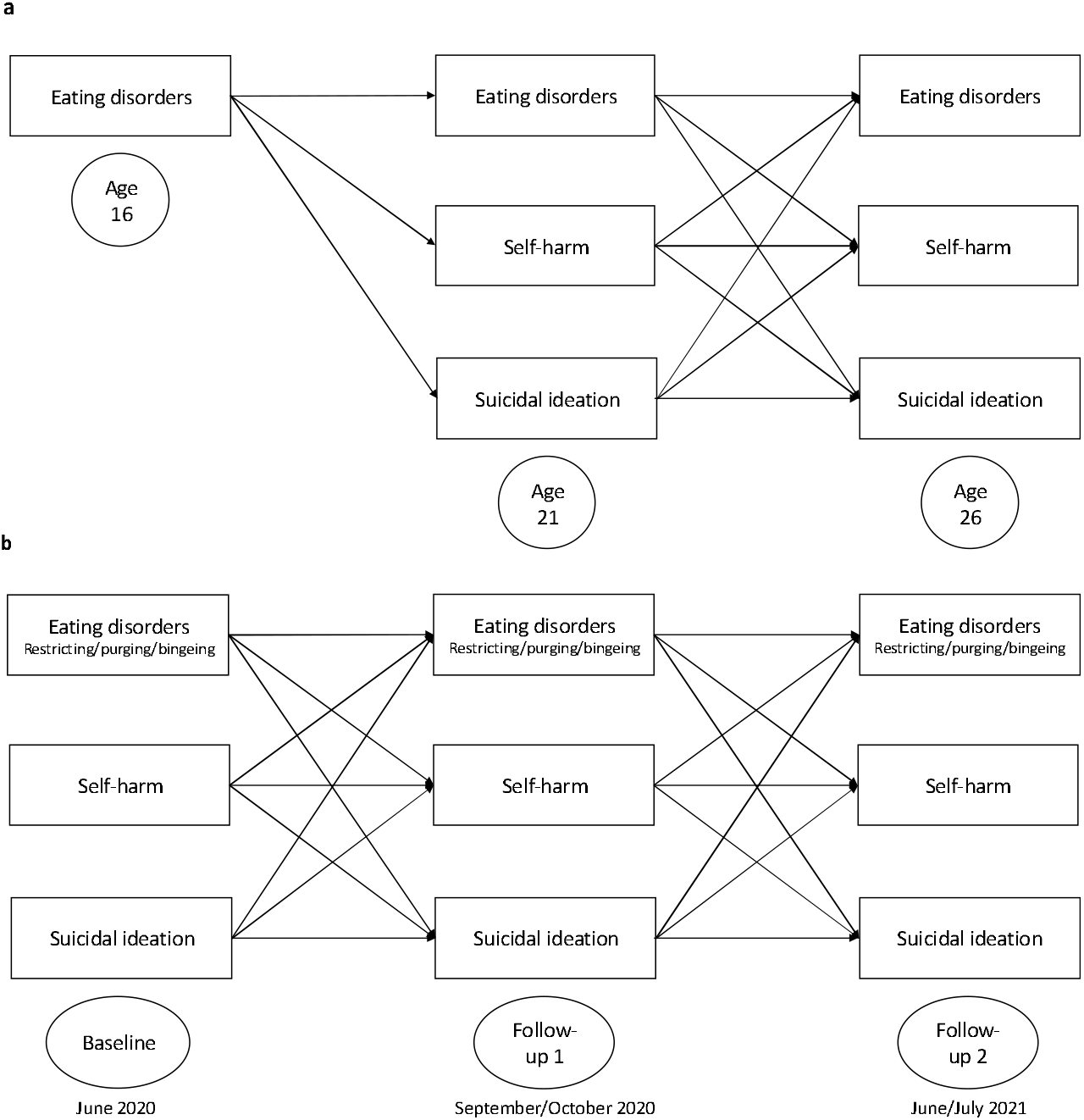
The longitudinal path diagrams. The models present the autoregressive longitudinal paths between symptoms of eating disorders, self-harm and suicidal ideation, as well as cross-trait longitudinal paths between the variables in TEDS (a) and COPING (b) samples.

## Results

### Longitudinal models in TEDS

In the TEDS cohort, the most severe symptoms of eating disorders, self-harm and suicidal ideation were on average reported when the twins were approximately 21 years of age and were subsequently observed to drop by the time the twins turned 26 (Figure 2a). At age 16, the eating disorder scores were significantly lower, compared to age 21 and 26 (Figure 2a). Results of the longitudinal SEM analysis are presented in Figure 3 for the total TEDS sample and Supplementary Figure 1 for the proportion of TEDS participants diagnosed with any eating disorder. Model fit indices and all path estimates with 95% confidence intervals (CIs) are included in Supplementary Tables 3 and 4. Eating disorders at age 16 were significantly associated with eating disorders at age 21 (β=0.51 [0.45, 0.56]), which in turn were significantly associated with eating disorders at age 26 (β=0.33 [0.29, 0.38]), demonstrating the continuity over time. Similarly, self-harm at age 21 demonstrated a significant relationship with self-harm at age 26 (β=0.11 [0.07, 0.16]), while suicidal ideation at age 21 was significantly associated with suicidal ideation at age 26 (β=0.29 [0.24, 0.34]), indicating stability across the two time points.

**Figure 2.**
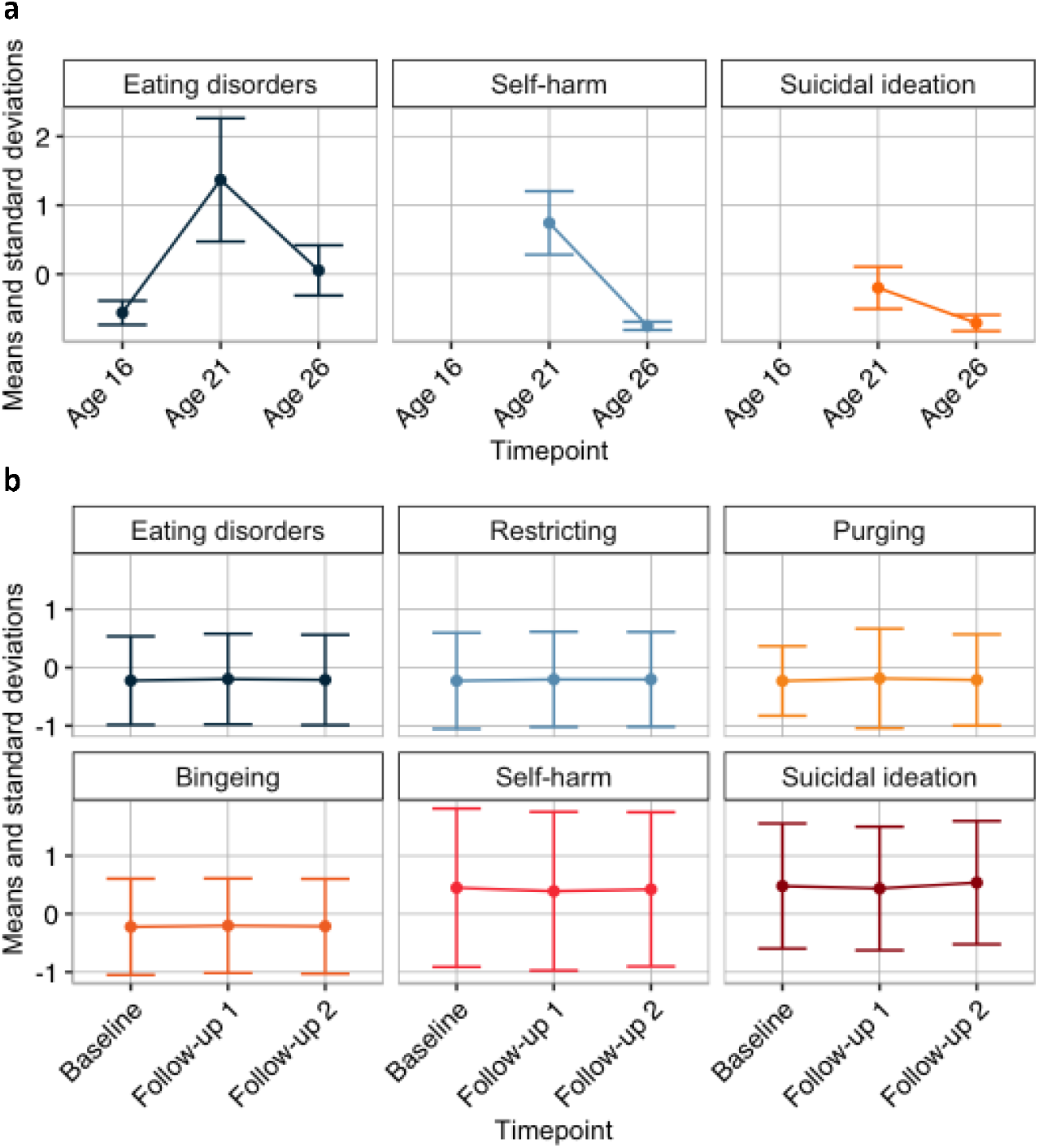
Means and standard deviations of the measures of eating disorders, self-harm and suicidal ideation in TEDS (a) and COPING (b) samples.

**Figure 3.**
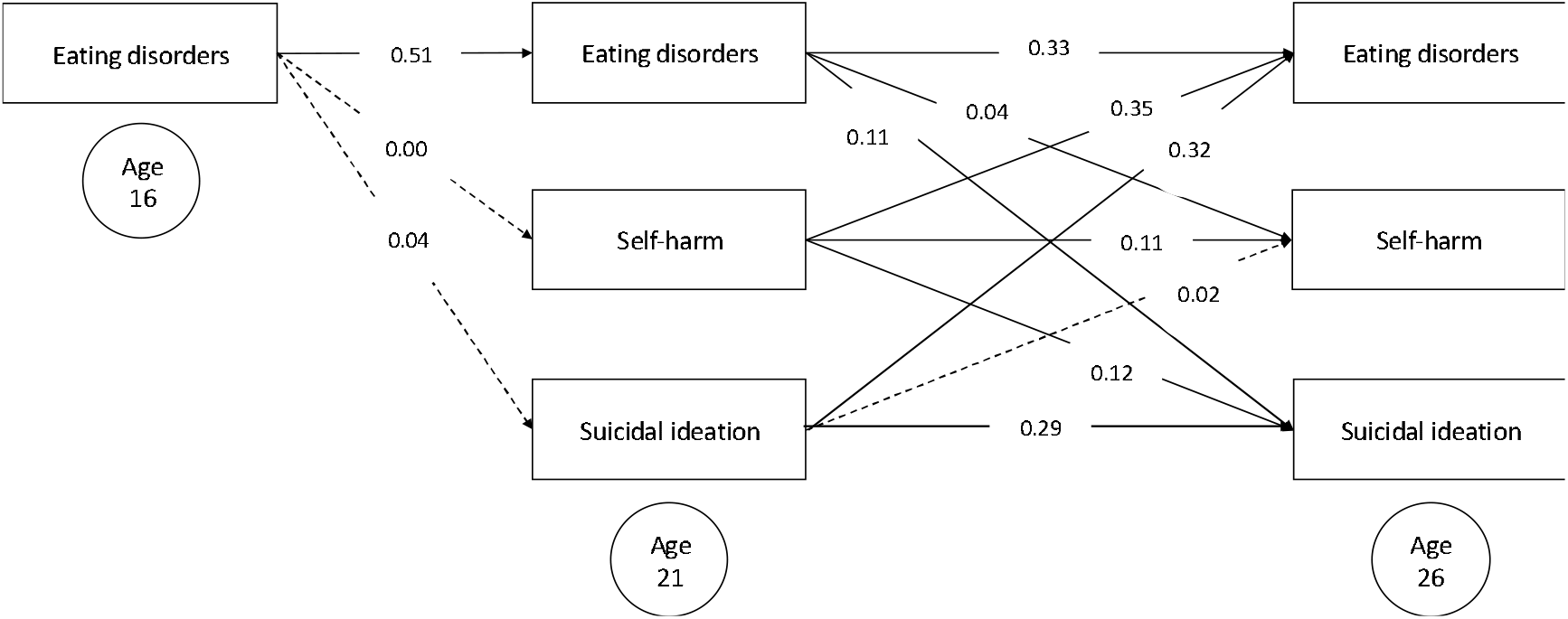
The longitudinal path diagram of the TEDS sample (*N*=5,196). The models present the autoregressive longitudinal paths between symptoms of eating disorders, self-harm and suicidal ideation, as well as cross-trait longitudinal paths between the variables.

Cross-domain associations showed more nuanced patterns. Eating disorders at age 16 did not show a significant effect on self-harm or suicidal ideation at age 21. However, eating disorders at age 21 were significantly associated with self-harm at age 26 (β=0.04, [ 0.01, 0.07]) and suicidal ideation at age 26 (β=0.11 [0.07, 0.14]). Notably, self-harm at age 21 was a significantly stronger predictor of eating disorders (β=0.35 [0.27, 0.43]), compared to suicidal ideation (β=0.12 [0.07, 0.17]) at age 26. Interestingly, suicidal ideation at age 21 were also strongly correlated with eating disorders at age 26 (β=0.32 [0.24, 0.40]) but not with self-harm at age 26. When the sample was restricted to those individuals who have been diagnosed with any eating disorder, only within-domain paths remained significant.

#### Longitudinal models in COPING

In the COPING cohort, the scores on general, as well as specific eating disorder measures, self-harm and suicidal ideation were nearly equivalent across the three timepoints (Figure 2b). Results of the SEM analysis are presented in Figure 4 for the total COPING sample and Supplementary Figure 2 for the proportion of COPING participants diagnosed with any eating disorder. The SEM results indicated that eating disorder symptoms exhibited moderate stability over time, with baseline eating disorders being significantly linked with eating disorders at follow-up 1 (β=0.45 [0.38, 0.52]), and follow-up 1 eating disorders with eating disorders at follow-up 2 (β=0.50 [0.43, 0.57]). The stability of restricting behaviours was higher, with baseline restricting being more strongly associated with follow-up 1 restricting (β=0.70 [0.65, 0.75]) and follow-up 1 restricting being strongly associated with follow-up 2 restricting (β=0.79 [0.76, 0.83]). Purging and bingeing behaviours also showed stability, although the effects were slightly lower than for restricting (purging: baseline to follow-up 1, β=0.64, [0.59, 0.69]; bingeing: baseline to follow-up 1, β=0.61, [ 0.56, 0.67]). Self-harm and suicidal ideation exhibited strong temporal stability across all models. Baseline self-harm demonstrated a strong relationship with follow-up 1 self-harm (β=0.54–0.56 across models), and follow-up 1 self-harm with follow-up 2 self-harm (β=0.59–0.60). Similarly, baseline suicidal ideation was linked to follow-up 1 suicidal ideation (β=0.60–0.61), and follow-up 1 suicidal ideation to follow-up 2 suicidal ideation (β=0.64 in all models).

**Figure 4.**
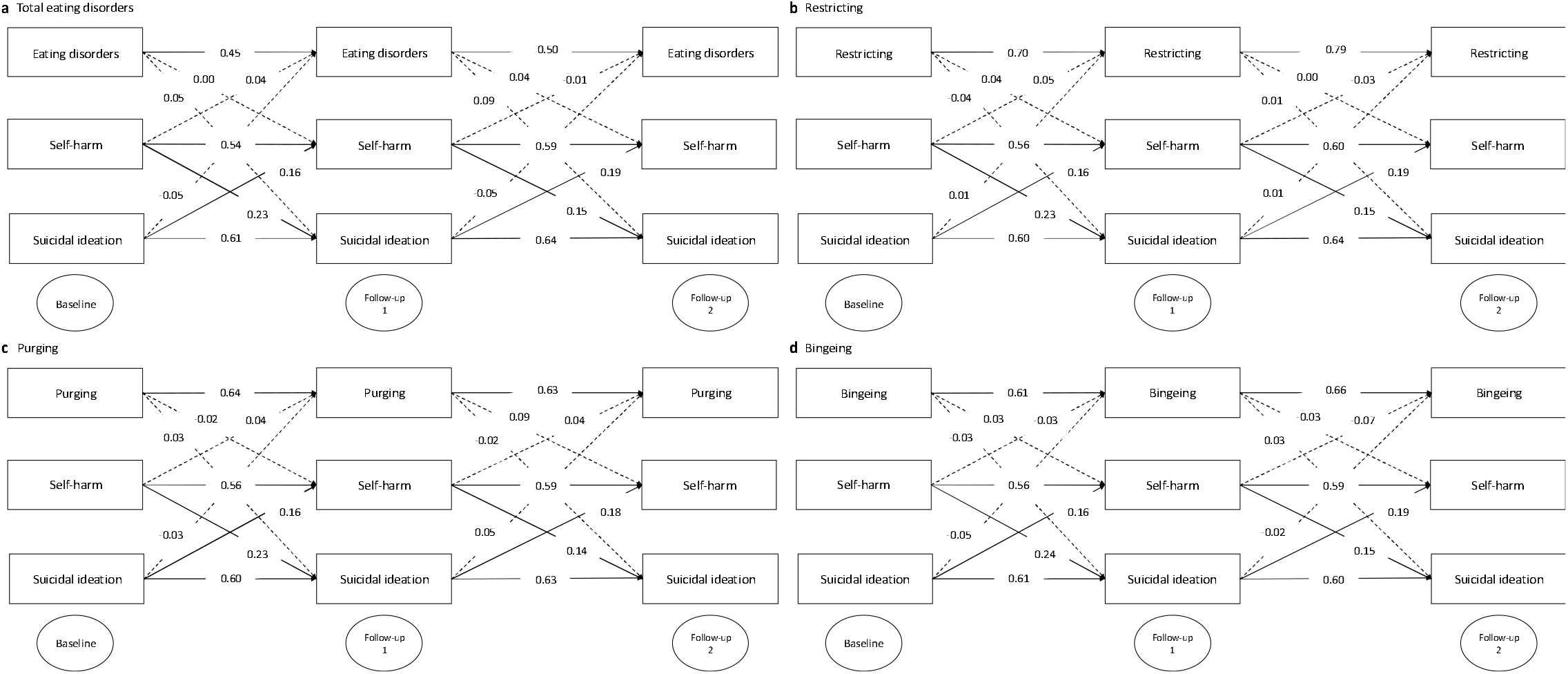
The longitudinal path diagrams of the COPING sample. The models present the autoregressive longitudinal paths between symptoms of eating disorders, self-harm and suicidal ideation, as well as cross-trait longitudinal paths between the variables in the total COPING sample (*N*=490). Results are presented for symptoms of any eating disorder (a), restricting (b), purging (c) and bingeing (d).

We found minimal evidence for cross-domain effects in COPING. Total eating disorders at baseline did not show a significant relationship with follow-up 1 self-harm or suicidal ideation. Similarly, baseline restricting, purging, and bingeing behaviours showed no significant associations with follow- up self-harm or suicidal ideation. However, follow-up 1 self-harm was significantly associated with follow-up 2 suicidal ideation in all models, with coefficients ranging from β=0.14 to 0.15.

## Discussion

This study examined the longitudinal relationships between eating disorders, self-harm, and suicidal ideation in two cohorts, TEDS and COPING, revealing distinct patterns of stability and cross-domain associations over time. In the TEDS cohort, symptoms of eating disorders, self-harm, and suicidal ideation demonstrated little continuity across adolescence and young adulthood, with peak severity observed around age 21 and decline by age 26. Cross-domain associations indicated that self-harm and suicidal ideation at age 26 were more strongly associated with symptoms of eating disorders at age 21 than vice versa. In contrast, the COPING cohort showed greater stability in symptoms over time but minimal evidence of cross-domain effects. In the TEDS cohort, symptoms of eating disorders were observed to peak at age 21, with a notable decline by age 26. This pattern suggests that while eating disorders may be most pronounced in young adulthood, there is a tendency for symptoms to decrease as individuals move into later stages of adulthood. Similar developmental trends were observed for symptoms of self-harm and suicidal ideation, which aligns with previous research indicating that self-harm may fluctuate or diminish as individuals age ^40^, perhaps in part because of the decrease in symptoms of eating disorders.

The finding that self-harm and suicidal ideation at age 21 were much stronger predictors of eating disorders at age 26 than the reverse suggests a directional relationship in which self-injurious thoughts and behaviours play a more central role in the development or persistence of disordered eating, rather than vice versa. This could indicate that self-harm and suicidal ideation represent earlier manifestations of underlying psychological distress, which later evolve into eating disorder symptoms as individuals seek alternative methods of emotional regulation, stress reduction or self-punishment ^41,42^. The markedly stronger association in this direction may also reflect the way in which self-harm functions as a coping mechanism for distress, with eating disorder behaviours emerging subsequently as an extension of these maladaptive strategies or as a more socially acceptable form of self-injurious behaviour ^43^. In contrast, the weaker effect of eating disorders on later self-harm or suicidal ideation suggests that, while these conditions often co-occur, eating pathology alone may not be as potent as risk factor for self-harm as self-harm is for eating disorders. Our findings align with the literature’s mixed results regarding the bidirectional relationship between eating disorders and suicidality. The observation that self-harm and suicidal ideation at age 21 are strongly associated with eating disorders at age 26 emphasises the pathway where self-harm precedes disordered eating ^11,12^.

The observed directional relationship where self-harm and suicidal ideation at age 21 are linked to eating disorder symptoms at age 26, aligns with a hypothesis of the overlap between symptoms of eating disorders and self-injury ^44^. This theory suggests that for some individuals, disordered eating behaviours—such as restricting, bingeing, purging, or excessive exercise—may serve as a method of self-harm rather than solely being driven by weight or shape concerns ^44,45^. Additionally, research indicating that nearly one-third of patients in intensive treatment for self-harm engage in disordered eating behaviours as a form of self-injury, reinforces the hypothesis that self-harm can manifest in multiple ways ^46^. The higher clinical severity at baseline among patients with self-injurious disordered eating behaviours mirrors the TEDS cohort’s findings, where self-harm and suicidality at an earlier stage were associated with later eating disorder symptoms, suggesting that self-injurious behaviours may evolve over time into different forms of psychological distress ^44^. The literature addressing the relationship between eating disorders and suicidality has been characterised by the lack of longitudinal studies ^9^. Most research in this area, including meta-analyses, has relied on cross-sectional and retrospective data, which cannot determine the directionality of the relationship between these two constructs. The only meta-analysis that has focused solely on longitudinal studies with suicide outcomes highlighted a substantial gap in this research, with only 14 longitudinal studies in the past 50 years exploring whether symptoms and diagnoses of eating disorders predict suicide attempts or death ^10^. Notably, no studies addressed suicidal ideation. This meta-analysis found that eating disorders were weak but significant predictors of suicide attempts, but not death ^10^.

In contrast, in the COPING cohort eating disorder symptoms demonstrated greater stability over time. This stability was evident in restrictive behaviours, which showed the strongest temporal associations, compared to purging and bingeing. Unlike the TEDS sample, cross-domain effects in COPING were generally weak, though self-harm and suicidal ideation exhibited strong reciprocal associations over time. This difference could be attributed to the unique clinical and demographic characteristics of the COPING sample. TEDS, as a representative population-based sample, captures a broad range of eating disorder symptoms, self-harm, and suicidal ideation. In contrast, COPING, as a clinical cohort, comprises individuals with a history of mood and anxiety disorders. This suggests that, in a clinical population with heightened emotional distress, self-harm and suicidal ideation may operate as distinct processes, while eating disorders may function as independent maladaptive coping mechanisms.

Another key factor contributing to the differential outcomes between the TEDS and COPING samples is the age of participants and the study design. TEDS follows a developmental trajectory, with the current data collected from adolescence into early adulthood, a critical period for the onset and fluctuation of eating disorders, self-harm, and suicidal ideation. The dynamic interplay between these behaviours in TEDS suggests that adolescence and early adulthood may be particularly sensitive windows for their co-occurrence. In contrast, COPING is a cross-sectional study with a much older mean age (late forties), capturing a population in which these behaviours may have stabilized over time. The greater temporal stability of eating disorder symptoms, particularly restrictive behaviours, in COPING could reflect the chronic nature of these disorders in individuals with a history of mood and anxiety disorders, who may have long-established maladaptive coping mechanisms. Moreover, the stronger associations between self-harm and suicidal ideation in COPING suggest that, in an older clinical population, these behaviours may be more ingrained and operate as distinct but interconnected constructs, whereas in younger, general-population samples like TEDS, they may still be in flux and more influenced by other psychopathological processes, including eating disorders.

Another crucial difference between the TEDS and COPING samples is the timing of data collection. The COPING data were gathered during the COVID-19 pandemic and associated lockdowns, a period marked by heightened psychological distress, social isolation, and disruptions to healthcare access. These factors may have contributed to the strong stability of symptoms, as individuals with preexisting mental health conditions might have experienced exacerbations of their symptoms due to pandemic-related stressors. In contrast, the TEDS data were collected across a broader developmental span, outside the immediate context of the pandemic, allowing for a more naturalistic examination of how these symptoms evolve from adolescence to early adulthood. The lack of significant cross-domain associations in COPING may reflect the unique psychosocial conditions of the pandemic, where distress was widespread but may not have followed typical trajectories of mental health co-occurrence. Conversely, the TEDS findings suggest that in a general-population sample, eating disorders, self-harm, and suicidal ideation may be potentially influenced by developmental and environmental factors beyond acute stressors. These differences highlight the need to establish broader contextual influences, such as global crises, when interpreting findings on mental health trajectories.

The clinical emphasis on early intervention in self-harm aligns with recommendations by the NICE (NICE, 2017), which stress the importance of addressing co-occurring psychiatric conditions such as self-harm to mitigate risks of both suicidality and eating disorders. Intervening at the stage of self-harm may not only reduce the immediate risk of suicide but also prevent the later onset or escalation of eating disorders. Since self-harm is a well-documented precursor to suicide in the general population ^47,48^, identifying and treating self-injurious behaviours early and addressing deficits in emotional regulation and maladaptive coping strategies could help break the cycle of distress and maladaptive coping before it leads to more severe psychopathology. Additionally, given the emerging evidence that self-harm and suicidal ideation may be stronger predictors of later eating disorders than the reverse, addressing these behaviours proactively could interrupt the trajectory leading to disordered eating. Since the evidence from the TEDS cohorts suggests that adolescence and early adulthood represent critical risk windows for the emergence of both eating disorders and self-harm, research is necessary to determine the precise nature of this relationship and whether early intervention in teenage populations could mitigate long-term risk trajectories ^49^. From a clinical standpoint, the findings suggest that treatment strategies for individuals engaging in self-harm should incorporate screening for emerging eating disorder symptoms. This could include routine screening for symptoms of eating disorders in individuals presenting with suicidal thoughts and behaviours.

Research suggests that eating disorder patients with a history of self-harm experience less reduction in body dissatisfaction and drive for thinness following inpatient treatment, but this effect becomes non-significant when controlling for negative affect ^50^. This suggests that self-harm may not independently hinder treatment outcomes but instead operates through broader emotional distress, encompassing emotion dysregulation, impulsivity, and distress intolerance ^51,52^. Since negative affect plays a central role in the persistence of both self-harm and disordered eating, interventions should prioritize emotional regulation strategies to improve treatment outcomes and reduce the likelihood of long-term psychopathology ^50,53^. Although the present findings suggest that self-harm and suicidal ideation in adolescence may be key risk factors for eating disorders in early adulthood, lack of data on self-harm and suicidality at age 16 limits our understanding of whether these behaviours in mid-adolescence predict later eating disorder onset, making this a priority for future research.

## Limitations

While this study provides important insights into the relationship between eating disorders and suicidal ideation, several limitations must be acknowledged. One key limitation is the COPING sample being substantially smaller due to poor data coverage across waves of repeated data collection, limiting statistical power and the ability to detect significant effects. Additionally, the COPING data were collected during the COVID-19 pandemic, a period of heightened psychological distress that may have resulted in the increase of severity of eating disorder symptoms and self-harm in ways not accounted for in the analyses ^20^. The short time frame between COPING measurement points—only a few months—also restricted the ability to observe meaningful changes in eating disorder behaviours or suicidal ideation. Another limitation is that we did not examine the role of co-occurring personality disorders, particularly borderline personality disorder, in mediating the relationship between symptoms of eating disorders and suicidality ^54,55^. While we chose not to focus on this perspective due to concerns about stigma and the reductive nature of such explanations, it remains a potential confounding factor that future research should consider.

## Conclusions

In summary, the present study highlights distinct longitudinal patterns between eating disorders, self-harm, and suicidal ideation across two cohorts, emphasizing the importance of developmental and contextual factors. The TEDS findings suggest that these behaviours are more dynamic and interrelated in adolescence and young adulthood, whereas the COPING cohort demonstrated greater temporal stability, particularly in restrictive eating behaviours. The stronger effects of self-harm and suicidal ideation on later eating disorders in TEDS highlight the necessity for targeted early support for suicidal thoughts and behaviours may help mitigate subsequent symptoms of eating disorders in young adulthood.

## Acknowledgements

We gratefully acknowledge the participation of all National Institute for Health and Care Research (NIHR) BioResource, the NIHR BioResource Centre Maudsley, Biomedical Research Centre at South London and Maudsley NHS Foundation Trust and King’s College London volunteers and thank the BioResource staff for their help with volunteer recruitment. We thank the NIHR Biomedical Research Centre at South London and the Maudsley NHS Foundation Trust and King’s College London for funding. We thank all participants who have kindly taken part in this study. This study represents independent research supported by the NIHR Biomedical Research Centre BioResource at South London and Maudsley NHS Foundation Trust and King’s College London. The views expressed in this publication are those of the authors and not necessarily those of the National Health Service, the NIHR or the UK Department of Health.

## Author contribution

AM and MH conceived and designed the study. AM analysed the data. AM wrote the paper with helpful contributions from UF, SK, TJ, EJT, RD, US, GB and MH. All authors contributed to the interpretation of the data, provided critical feedback on paper drafts and approved the final draft.

## Transparency Declaration

The manuscript is an honest, accurate, and transparent account of the study being reported. No important aspects of the study have been omitted and any discrepancies from the study as planned and pre-registered have been explained.

## Data Availability

Data availability is not applicable to this article as no new data were created or analysed in this study.

## Analytic Code Availability

Analytic scripts are available via https://github.com/agmusial/longitudinal_links_eds_sh_si.

