## Supplementary Material for "Reciprocal longitudinal associations between symptoms of eating disorders, self-harm and suicidal ideation"

Supplementary Notes

Supplementary Note 1

Hypotheses pre-registered with the Open Science Framework (OSF).

We have two hypotheses based on previous literature.

1.	Within-variable longitudinal associations will be stronger than between-variable associations across timepoints, i.e., eating disorder symptoms at Time 1 will more strongly predict eating disorder symptoms as Time 2 and Time 3, compared to the prediction of subsequent self-harm and suicidal ideation.

2.	The longitudinal prediction between symptoms of eating disorders and suicidal ideation will be mediated by self-harm.

Supplementary Note 2

The role of psychosocial factors in the relationship between eating disorder and suicidality.

*Background*

Limited research has explored whether the link between eating disorders and self-harm behaviours is influenced by other psychosocial factors. Individuals with eating disorders might experience profound feelings of loneliness and social isolation, which can intensify symptom severity, thereby increasing their vulnerability to suicidal ideation ^1–3^. Conversely, positive interpersonal relationships can provide emotional support, reduce feelings of loneliness, and improve overall mental health, including improved eating disorder symptom severity ^4^. Other relevant psychosocial factors that influence the risk for suicidality in individuals experiencing symptoms of eating disorders include substance abuse ^5,6^ and salient life events, such as parental divorce, serious illness or moving houses ^7,8^.

*Psychosocial variables*

In TEDS, psychosocial variables included measures of relationships with siblings and parents (Fraley et al., 2011; Riggio, 2000), life satisfaction ^11^, health behaviours ^12^ and the count of life events, which included circumstances like financial difficulties, serious illness or divorce ^13^. In COPING, we included measures of loneliness, grief, health behaviours and incidence of negative life events. In TEDS, those measures were collected when the twins were approximately 21 years of age, while in COPING the data were collected in June 2020, following the baseline data collection. For a list of scales and items please refer to Supplementary Tables 1 and 2.

*Mediation models*

We performed the mediation analyses between symptoms of eating disorders at baseline and suicidal ideation at the final timepoint (Supplementary Figure 3). The candidate mediators included mid-point measures of psychosocial variables related to interpersonal relationships, health behaviours and life events. Mediators were selected based on their correlations with predictor and outcome variables. We did not set the correlation threshold but rather selected measures that were most strongly correlated with baseline eating disorders and final measurement of suicidal ideation. In TEDS, the mediation models did not differentiate between different sub-types of eating disorders, whereas in COPING, we distinguished between symptoms related to restricting, purging and bingeing. To achieve that, we initially ran separate mediation models for the three eating disorder sub-types and subsequently refitted the models collapsing the groups and compared the model fit indices between the models that do and do not distinguish between groups.

*Results*

Upon investigation of the correlation matrices in TEDS, the psychosocial variables to be tested as mediators included, love and relationships, community, relationship with twin and count of difficult life events, based on the correlations with both the predictor and the outcome being larger or equal to 0.05 or –0.05 (Supplementary Figure 4a). In the first model**, love and relationships** did not significantly mediate the relationship between **eating disorders at age 16** and **suicidal ideation at age 26** (ab=0.01, p=0.38), with the total effect primarily driven by the direct effect (c=0.09, p=0.002). In the second model, **community support** also showed no significant mediation (ab=0.01, p=0.12), though the total effect (c=0.08, p=0.01) remained significant. The third analysis revealed partial mediation via **the relationship with twin variable** (ab=0.01, p=0.06), with a significant total effect (c=0.10, p=0.001). Finally, the fourth model demonstrated significant mediation by **life events** (ab=0.02, p=0.001), accounting for a substantial portion of the total effect (c=0.09, p=0.003). Overall, **the count of difficult life events** emerged as the most robust mediator, with the indirect effects accounting for 26% of the total effect, while other mediators had limited contributions.

In the COPING sample, the selected mediators included loneliness and relationship loss-related grief for all analyses (Supplementary Figures 4b-e). The analysis involving restricting additionally included positive health behaviours as potential mediator and analyses involving purging and bingeing as predictors additionally included both positive and negative health behaviours (Supplementary Figures 6c-e). While the total effect of **eating disorder symptoms at baseline on suicidal ideation at the final follow-up w**as significant (c=0.25, p=0.002), the indirect effect via loneliness was nonsignificant (ab=0.02, p=0.62). Similarly, the indirect effect of relationship loss-related grief was negligible and nonsignificant (ab=0.002, p=0.81), suggesting no evidence of significant indirect pathways through the tested mediators. No significant mediation effects were observed in the analyses of restricting, purging and bingeing. Full results of the mediation analyses are presented in Supplementary Table 6.

*Conclusions*

The mediation analyses provided additional exploratory insights into the factors that may explain the relationship between eating disorders and suicidal ideation. In the TEDS cohort, the count of difficult life events emerged as a robust mediator, accounting for 26% of the total effect of symptoms of eating disorders at age 16 and suicidal ideation at age 26. These findings suggest that the accumulation of negative life events, such as experiences of trauma, loss, or significant stress, may play a critical role in exacerbating the effects of eating disorders on suicidal ideation over time. The lack of significant mediation in the analyses of restricting, purging, and bingeing behaviours in the COPING sample supports the idea that the relationship between eating disorder symptoms and suicidal ideation may be more individualized and influenced by other variables not measured in the present study, such as experiences related to treatment.

Supplementary Tables

Supplementary Table 1. List of items in TEDS.

Eating disorder items

Age 16

Have you felt fat?

Have you had a definite fear that you might gain weight or become fat?

Has your weight influenced how you think about yourself as a person?

Has your shape influenced how you think about yourself as a person?

Age 21

I stuff myself with food.

I think about dieting.

I am terrified of gaining weight.

I am preoccupied with the desire to be thinner.

I think about bingeing (overeating).

I think my hips are too big.

If I gain a pound, I worry that I will keep gaining.

I have the thought of trying to vomit in order to lose weight.

I think my buttocks are too large.

I eat or drink in secrecy.

Age 26

Have you ever had a period of time when you weighed much less than other people thought you should weigh?

Have you ever had recurrent episodes of excessive overeating or binge eating (i.e., eating significantly more than what most people eat in a similar period of time, for example, 2 hours)?

During your episodes of excessive overeating/binge eating, how often did you feel like you had no control over your eating (e.g., not being able to stop eating or feeling compelled to eat)?

During these episodes of excessive overeating/binge eating, did you: Eat much more rapidly than normal; Eat until feeling uncomfortably full; Eat large amounts of food when not feeling physically hungry; Eat alone because of feeling embarrassed by how much you are eating; Feel disgusted, depressed, or very guilty afterward?

Do you feel distressed about your episodes of excessive overeating/binge eating?

To compensate for overeating, have you used any of the following at least once a week for at least 3 months? Fasted for 8 waking hours or longer; Made yourself vomit; Used diet pills, laxatives, diuretics, drugs; Exercised excessively or compulsively

In general, how dependent has your self-esteem been on your body shape or weight?

Independent from low weight or excessive overeating, have you used any of the following at least once a week for at least 3 months, to control your weight or shape? Fasted for 8 waking hours or longer; Made yourself vomit; Used diet pills, laxatives, diuretics, drugs; Exercised excessively or compulsively?

Self-harm items

Age 21

In your lifetime, have you ever hurt yourself on purpose in any way (e.g. by taking an overdose of pills, or by cutting yourself?

In your lifetime, have you ever hurt yourself on purpose without intending to kill yourself?

In your lifetime, did you hurt yourself because you wanted to show how desperate you were feeling?

In your lifetime, did you hurt yourself because you wanted to punish yourself?

In your lifetime, did you hurt yourself because you wanted to frighten someone?

In your lifetime, did you hurt yourself ecause you wanted to get relief from a terrible state of mind?

After hurting yourself on purpose, have you ever sought medical help/first aid from your GP (family doctor)?

After hurting yourself on purpose, have you ever sought medical help/first aid from hospital casualty/ emergency department?

After hurting yourself on purpose, have you ever sought medical help/first aid from another healthcare professional?

Age 26

In the past year, have you ever hurt or harmed yourself on purpose in any way (e.g., by taking an overdose of pills, or by cutting yourself)?

Suicidal ideation items

Age 21

In your lifetime, have you ever thought about killing yourself, even if you would not really do it?

In your lifetime, on any of the occasions you have hurt yourself on purpose, have you ever seriously wanted to kill yourself?

In your lifetime, did you hurt yourself ecause you wanted to die?

Age 26

In the past year, have you ever thought about killing yourself, even if you would not really do it?

Psychosocial measures

Relationship with twin

I enjoy my relationship with my twin.

My twin and I have a lot of fun together.

I like to spend time with my twin.

My twin and I do a lot of things together.

My twin talks to me about personal problems.

Relationship with mother

It helps to turn to my mother in times of need.

I usually discuss my problems and concerns with my mother.

I talk things over with my mother.

I find it easy to depend on my mother.

I don't feel comfortable opening up to my mother.

Relationship with father

It helps to turn to my father in times of need.

I usually discuss my problems and concerns with my father.

I talk things over with my father.

I find it easy to depend on my father.

I don't feel comfortable opening up to my father.

Contentment with life assessment scale (CLAS): love and relationships

I am happy with my love life.

I have the level of intimacy in my life that I want.

I feel loved.

Contentment with life assessment scale (CLAS): community satisfaction

I enjoy living in my neighbourhood.

I would prefer to move to a different area if I could.

I feel a sense of belonging in my neighbourhood.

I feel like an outsider in my neighbourhood.

I feel safe in my neighbourhood.

Rapid Eating and Activity Assessment for Patients (REAP)

How often do you eat 3 portions of whole grain products in one day? (e.g. brown rice, brown bread).

How often do you eat 5 portions of fruit and vegetables in one day?

How often do you eat 3-4 portions of milk and dairy foods or dairy alternatives in one day?

How often do you eat processed meats/fish? (skip this question if you are vegetarian/vegan).

How often do you eat 2 portions of protein-rich foods in one day, including fish, white meat, beans/pulses, soya based products?

How often do you eat fried foods such as fried chicken, fried fish or chips?

How often do you eat salted snacks such as crisps, crackers, nachos, etc.?

How often do you eat unsalted snacks such as nuts, seeds, rice crackers, air-popped popcorn, etc.?

How often do you use oils such as olive, rapeseed, sunflower, flaxseed, etc.?

How often do you eat sweets like cake, cookies, pastries, chocolate, ice cream, muffins, etc.?

How often do you drink 1 can or more of fizzy drinks or fruit drinks made from concentrate in one day?

How often do you eat processed foods like canned soup, frozen/packaged meals, chips?

Life events

Since you were 16 years of age you became homeless.

Since you were 16 years of age you or your partner became pregnant or had a baby.

Since you were 16 years of age you lost your job or got into serious financial problems.

Since you were 16 years of age you were divorced or separated.

Since you were 16 years of age you were admitted to hospital or became seriously ill.

Since you were 16 years of age you were in trouble with the law.

Since you were 16 years of age you were the victim of a serious crime.

Since you were 16 years of age someone close to you died.

Since you were 16 years of age you attempted suicide.

Since you were 16 years of age you or your partner had an abortion.

Since you were 16 years of age your parents divorced.

Supplementary Table 2. List of items in COPING.

Eating disorder items

On how many of the past 28 days have you been deliberately trying to limit the amount of food you eat to influence your shape or weight?

On how many of the past 28 days has thinking about body shape, weight, food, eating, or calories made it very difficult to concentrate on things you are interested in?

Over the past 28 days has your weight influenced how you think about yourself as a person?

Over the past 28 days how dissatisfied have you been with your weight or shape?

Over the past 28 days how many times per day have you actively been monitoring your shape or weight?

Over the past 28 days how many times per day have you eaten what other people would regard as an unusually large amount of food with a sense of having lost control over your eating?

Over the past 28 days how many times per day have you made yourself sick as a means of controlling your shape or weight?

Over the past 28 days how many times per day have you taken laxatives, diuretics, or diet pills as a means of controlling your shape or weight?

Over the past 28 days how many times per day have you exercised in a “driven” or “compulsive” way as a means of controlling your weight, shape, or amount of fat, or to burn off calories?

Over the past 28 days how distressed have you been by your eating behaviours, the measures you've taken to control your weight, or concerns about your weight and shape?

Over the past 28 days, on how many days have you picked at, nibbled, or grazed food repeatedly outside of planned meals and snacks?

Over the past 28 days, on how many days have you eaten more than usual because you felt anxious, blue, bored, frustrated or lonely?

Self-harm items

Have you contemplated harming yourself?

Before the pandemic, had you deliberately harmed yourself, whether or not you meant to end your life?

Suicidal ideation item

Many people have thoughts that life is not worth living. Have you felt that way?

Psychosocial measures

Loneliness

Over the past two weeks, how often have you felt that you lack companionship?

Over the past two weeks, how often have you felt left out?

Over the past two weeks, how often have you felt isolated from others?

Over the past two weeks, how often have you felt alone?

Grief: Covid-19 pandemic

In the past month, have you lost someone close to you due to COVID-19?

Grief: relationships

In the past month, have you suffered the loss of any significant relationship?

Self-care: positive

Over the past two weeks, how often have you got enough sleep?

Over the past two weeks, how often have you eaten nutritiously?

Over the past two weeks, how often have you exercised?

Over the past two weeks, how often have you stayed hydrated?

Over the past two weeks, how often have you taken extra vitamins or supplements?

Over the past two weeks, how often have you taken part in leisure activities you enjoy?

Over the past two weeks, how often have you maintained normal levels of hygiene?

Over the past two weeks, how often have you maintained your normal daily structure?

Over the past two weeks, how often have you socialised with people inside your home?

Over the past two weeks, how often have you socialised with people outside your home?

Over the past two weeks, how often have you socialised with people outside your home virtually?

Over the past two weeks, how often have you socialised with people outside your home in person?

Over the past two weeks, how often have you meditated or used other relaxation techniques?

Over the past two weeks, how often have you prayed or other spiritual activities?

Over the past two weeks, how often have you spent time in a private outdoor space?

Over the past two weeks, how often have you spent time in public outdoor space?

Self-care: negative

Over the past two weeks, how often have you smoked cigarettes or vaped?

Over the past two weeks, how often have you smoked cannabis?

Over the past two weeks, how often have you drunk alcohol?

Over the past two weeks, how often have you used other recreational drugs?

Life events

In the last two weeks, have you experienced any major life events unrelated to the pandemic that you feel have affected any of your answers? What was the nature of this life event/events?

Supplementary Table 3. Model fit indices.

| Model | CFI | TLI | AIC | BIC | RMSEA (95% CIs) |
| --- | --- | --- | --- | --- | --- |
| TEDS | | | | | |
| Total sample | 0.551 | -0.094 | 85546.803 | 85705.568 | 0.122 (0.117, 0.127) |
| Individuals diagnosed with an eating disorder | 0.587 | -0.007 | 4270.358 | 4275.300 | 0.141 (0.119, 0.165) |
| COPING | | | | | |
| Total sample | | | | | |
| Total eating disorders | 0.801 | 0.285 | 12414.069 | 12472.234 | 0.241 (0.221, 0.260) |
| Restricting | 0.836 | 0.535 | 11953.675 | 12007.757 | 0.222 (0.205, 0.240) |
| Purging | 0.792 | 0.409 | 12062.622 | 12116.705 | 0.233 (0.216, 0.251) |
| Bingeing | 0.822 | 0.494 | 12336.781 | 12390.864 | 0.214 (0.197, 0.231) |
| Individuals diagnosed with an eating disorder | | | | | |
| Total eating disorders | 0.783 | 0.219 | 2494.469 | 2455.172 | 0.223 (0.117, 0.273) |
| Restricting | 0.833 | 0.527 | 2180.551 | 2144.012 | 0.213 (0.172, 0.257) |
| Purging | 0.756 | 0.307 | 2541.542 | 2505.002 | 0.240 (0.198, 0.283) |
| Bingeing | 0.781 | 0.377 | 2342.448 | 2305.909 | 0.243 (0.202, 0.287) |

*Note*. CFI= comparative fit index; TLI= Tucker-Lewis Index; AIC= Akaike information criterion; BIC= Bayesian information criterion; RMSEA= Root Mean Square Error of Approximation; CIs= confidence intervals.

Supplementary Table 4. Path estimates and 95% confidence intervals.

| Model | Path | Standardized estimate and 95% CIs |
| --- | --- | --- |
| TEDS | | |
| Total sample | Age 16 EDs ~ Age 21 EDs | 0.505 (0.449, 0.561) |
|  | Age 21 EDs ~ Age 26 EDs | 0.334 (0.292, 0.377) |
|  | Age 21 SH ~ Age 26 SH | 0.114 (0.073, 0.156) |
|  | Age 21 SI ~ Age 26 SI | 0.289 (0.239, 0.338) |
|  | Age 16 EDs ~ Age 21 SH | 0.00 (-0.088, 0.087) |
|  | Age 16 EDs ~ Age 21 SI | 0.038 (-0.051, 0.126) |
|  | Age 21 EDs ~ Age 26 SH | 0.039 (0.007, 0.071) |
|  | Age 21 EDs ~ Age 26 SI | 0.106 (0.073, 0.139) |
|  | Age 21 SH ~ Age 26 EDs | 0.350 (0.266, 0.433) |
|  | Age 21 SH ~ Age 26 SI | 0.117 (0.066, 0.167) |
|  | Age 21 SI ~ Age 26 EDs | 0.322 (0.241, 0.404) |
|  | Age 21 SI ~ Age 26 SH | 0.024 (-0.017, 0.065) |
| Individuals diagnosed with an eating disorder | Age 16 EDs ~ Age 21 EDs | 0.581 (0.313, 0.849) |
|  | Age 21 EDs ~ Age 26 EDs | 0.292 (0.039, 0.545) |
|  | Age 21 SH ~ Age 26 SH | 0.233 (0.102, 0.363) |
|  | Age 21 SI ~ Age 26 SI | 0.240 (0.042, 0.437) |
|  | Age 16 EDs ~ Age 21 SH | 0.222 (-0.252, 0.696) |
|  | Age 16 EDs ~ Age 21 SI | -0.121 (-0.380, 0.139) |
|  | Age 21 EDs ~ Age 26 SH | 0.121 (-0.014, 0.255) |
|  | Age 21 EDs ~ Age 26 SI | 0.091 (-0.076, 0.257) |
|  | Age 21 SH ~ Age 26 EDs | 0.108 (-0.312, 0.527) |
|  | Age 21 SH ~ Age 26 SI | 0.192 (-0.001, 0.385) |
|  | Age 21 SI ~ Age 26 EDs | -0.064 (-0.507, 0.380) |
|  | Age 21 SI ~ Age 26 SH | 0.129 (-0.066, 0.324) |
| COPING | | |
| Total sample | Baseline EDs ~ Follow-up 1 EDs | 0.454 (0.383, 0.524) |
|  | Follow-up 1 EDs ~ Follow-up 2 EDs | 0.502 (0.434, 0.569) |
|  | Baseline SH ~ Follow-up 1 SH | 0.543 (0.470, 0.616) |
|  | Follow-up 1 SH ~ Follow-up 2 SH | 0.594 (0.520, 0.669) |
|  | Baseline SI ~ Follow-up 1 SI | 0.612 (0.539, 0.685) |
|  | Follow-up 1 SI ~ Follow-up 2 SI | 0.638 (0.566, 0.711) |
|  | Baseline EDs ~ Follow-up 1 SH | -0.001 (-0.097, 0.094) |
|  | Baseline EDs ~ Follow-up 1 SI | 0.052 (-0.044, 0.147) |
|  | Follow-up 1 EDs ~ Follow-up 2 SH | 0.036 (-0.068, 0.141) |
|  | Follow-up 1 EDs ~ Follow-up 2 SI | 0.092 (-0.012, 0.196) |
|  | Baseline SH ~ Follow-up 1 EDs | 0.037 (-0.029, 0.103) |
|  | Baseline SH ~ Follow-up 1 SI | 0.225 (0.143, 0.308) |
|  | Follow-up 1 SH ~ Follow-up 2 EDs | -0.009 (-0.076, 0.059) |
|  | Follow-up 1 SH ~ Follow-up 2 SI | 0.150 (0.058, 0.242) |
|  | Baseline SI ~ Follow-up 1 EDs | -0.045 (-0.114, 0.025) |
|  | Baseline SI ~ Follow-up 1 SH | 0.161 (0.073, 0.248) |
|  | Follow-up 1 SI ~ Follow-up 2 EDs | -0.053 (-0.122, 0.017) |
|  | Follow-up 1 SI ~ Follow-up 2 SH | 0.189 (0.091, 0.288) |
| Total sample | Baseline restricting ~ Follow-up 1 restricting | 0.700 (0.653, 0.746) |
|  | Follow-up 1 restricting ~ Follow-up 2 restricting | 0.791 (0.756, 0.826) |
|  | Baseline SH ~ Follow-up 1 SH | 0.564 (0.493, 0.635) |
|  | Follow-up 1 SH ~ Follow-up 2 SH | 0.596 (0.521, 0.670) |
|  | Baseline SI ~ Follow-up 1 SI | 0.603 (0.529, 0.676) |
|  | Follow-up 1 SI ~ Follow-up 2 SI | 0.635 (0.561, 0.708) |
|  | Baseline restricting ~ Follow-up 1 SH | 0.039 (-0.035, 0.113) |
|  | Baseline restricting ~ Follow-up 1 SI | -0.036 (-0.108, 0.037) |
|  | Follow-up 1 restricting ~ Follow-up 2 SH | 0.000 (-0.073, 0.074) |
|  | Follow-up 1 restricting ~ Follow-up 2 SI | 0.009 (-0.068, 0.085) |
|  | Baseline SH ~ Follow-up 1 restricting | 0.048 (-0.020, 0.116) |
|  | Baseline SH ~ Follow-up 1 SI | 0.233 (0.150, 0.316) |
|  | Follow-up 1 SH ~ Follow-up 2 restricting | -0.029 (-0.099, 0.040) |
|  | Follow-up 1 SH ~ Follow-up 2 SI | 0.151 (0.058, 0.243) |
|  | Baseline SI ~ Follow-up 1 restricting | 0.013 (-0.062, 0.087) |
|  | Baseline SI ~ Follow-up 1 SH | 0.159 (0.072, 0.246) |
|  | Follow-up 1 SI ~ Follow-up 2 restricting | 0.008 (-0.064, 0.079) |
|  | Follow-up 1 SI ~ Follow-up 2 SH | 0.189 (0.090, 0.288) |
| Total sample | Baseline purging ~ Follow-up 1 purging | 0.638 (0.585, 0.692) |
|  | Follow-up 1 purging ~ Follow-up 2 purging | 0.629 (0.574, 0.683) |
|  | Baseline SH ~ Follow-up 1 SH | 0.561 (0.489, 0.632) |
|  | Follow-up 1 SH ~ Follow-up 2 SH | 0.593 (0.519, 0.668) |
|  | Baseline SI ~ Follow-up 1 SI | 0.604 (0.532, 0.677) |
|  | Follow-up 1 SI ~ Follow-up 2 SI | 0.632 (0.558, 0.705) |
|  | Baseline purging ~ Follow-up 1 SH | -0.016 (-0.100, 0.067) |
|  | Baseline purging ~ Follow-up 1 SI | 0.026 (-0.055, 0.106) |
|  | Follow-up 1 purging ~ Follow-up 2 SH | 0.087 (-0.006, 0.180) |
|  | Follow-up 1 purging ~ Follow-up 2 SI | -0.020 (-0.111, 0.072) |
|  | Baseline SH ~ Follow-up 1 purging | 0.040 (-0.028, 0.107) |
|  | Baseline SH ~ Follow-up 1 SI | 0.234 (0.152, 0.317) |
|  | Follow-up 1 SH ~ Follow-up 2 purging | 0.042 (-0.025, 0.110) |
|  | Follow-up 1 SH ~ Follow-up 2 SI | 0.141 (0.049, 0.234) |
|  | Baseline SI ~ Follow-up 1 purging | -0.029 (-0.098, 0.041) |
|  | Baseline SI ~ Follow-up 1 SH | 0.163 (0.076, 0.250) |
|  | Follow-up 1 SI ~ Follow-up 2 purging | 0.054 (-0.014, 0.123) |
|  | Follow-up 1 SI ~ Follow-up 2 SH | 0.182 (0.082, 0.281) |
| Total sample | Baseline bingeing ~ Follow-up 1 bingeing | 0.611 (0.555, 0.666) |
|  | Follow-up 1 bingeing ~ Follow-up 2 bingeing | 0.658 (0.606, 0.709) |
|  | Baseline SH ~ Follow-up 1 SH | 0.560 (0.489, 0.631) |
|  | Follow-up 1 SH ~ Follow-up 2 SH | 0.593 (0.518, 0.667) |
|  | Baseline SI ~ Follow-up 1 SI | 0.608 (0.535, 0.680) |
|  | Follow-up 1 SI ~ Follow-up 2 SI | 0.635 (0.562, 0.708) |
|  | Baseline bingeing ~ Follow-up 1 SH | 0.025 (-0.059, 0.109) |
|  | Baseline bingeing ~ Follow-up 1 SI | -0.031 (-0.114, 0.052) |
|  | Follow-up 1 bingeing ~ Follow-up 2 SH | -0.026 (-0.118, 0.066) |
|  | Follow-up 1 bingeing ~ Follow-up 2 SI | 0.029 (-0.063, 0.121) |
|  | Baseline SH ~ Follow-up 1 bingeing | -0.025 (-0.092, 0.041) |
|  | Baseline SH ~ Follow-up 1 SI | 0.243 (0.161, 0.325) |
|  | Follow-up 1 SH ~ Follow-up 2 bingeing | -0.070 (-0.136, -0.003) |
|  | Follow-up 1 SH ~ Follow-up 2 SI | 0.152 (0.060, 0.244) |
|  | Baseline SI ~ Follow-up 1 bingeing | -0.053 (-0.124, 0.018) |
|  | Baseline SI ~ Follow-up 1 SH | 0.155 (0.068, 0.241) |
|  | Follow-up 1 SI ~ Follow-up 2 bingeing | -0.024 (-0.093, 0.045) |
|  | Follow-up 1 SI ~ Follow-up 2 SH | 0.188 (0.089, 0.287) |
| Individuals diagnosed with an eating disorder | Baseline EDs ~ Follow-up 1 EDs | 0.365 (0.183, 0.547) |
|  | Follow-up 1 EDs ~ Follow-up 2 EDs | 0.409 (0.231, 0.586) |
|  | Baseline SH ~ Follow-up 1 SH | 0.579 (0.423, 0.736) |
|  | Follow-up 1 SH ~ Follow-up 2 SH | 0.548 (0.364, 0.732) |
|  | Baseline SI ~ Follow-up 1 SI | 0.731 (0.609, 0.853) |
|  | Follow-up 1 SI ~ Follow-up 2 SI | 0.691 (0.539, 0.842) |
|  | Baseline EDs ~ Follow-up 1 SH | 0.055 (-0.166, 0.276) |
|  | Baseline EDs ~ Follow-up 1 SI | 0.002 (-0.231, 0.234) |
|  | Follow-up 1 EDs ~ Follow-up 2 SH | 0.012 (-0.236, 0.261) - |
|  | Follow-up 1 EDs ~ Follow-up 2 SI | 0.054 (-0.308, 0.199) |
|  | Baseline SH ~ Follow-up 1 EDs | 0.047 (-0.110, 0.205) |
|  | Baseline SH ~ Follow-up 1 SI | 0.147 (-0.044, 0.337) |
|  | Follow-up 1 SH ~ Follow-up 2 EDs | 0.068 (-0.101, 0.237) |
|  | Follow-up 1 SH ~ Follow-up 2 SI | 0.184 (-0.039, 0.408) - |
|  | Baseline SI ~ Follow-up 1 EDs | 0.090 (-0.232, 0.051) |
|  | Baseline SI ~ Follow-up 1 SH | 0.018 (-0.157, 0.193) - |
|  | Follow-up 1 SI ~ Follow-up 2 EDs | 0.123 (-0.290, 0.044) |
|  | Follow-up 1 SI ~ Follow-up 2 SH | 0.083 (-0.132, 0.298) |
| Individuals diagnosed with an eating disorder | Baseline restricting ~ Follow-up 1 restricting | 0.815 (0.736, 0.895) |
|  | Follow-up 1 restricting ~ Follow-up 2 restricting | 0.746 (0.649, 0.843) |
|  | Baseline SH ~ Follow-up 1 SH | 0.587 (0.432, 0.743) |
|  | Follow-up 1 SH ~ Follow-up 2 SH | 0.554 (0.371, 0.736) |
|  | Baseline SI ~ Follow-up 1 SI | 0.741 (0.616, 0.866) |
|  | Follow-up 1 SI ~ Follow-up 2 SI | 0.706 (0.561, 0.851) |
|  | Baseline restricting ~ Follow-up 1 SH | 0.073 (-0.068, 0.215) |
|  | Baseline restricting ~ Follow-up 1 SI | -0.124 (-0.271, 0.023) |
|  | Follow-up 1 restricting ~ Follow-up 2 SH | -0.176 (-0.336, -0.016) |
|  | Follow-up 1 restricting ~ Follow-up 2 SI | 0.267 (0.097, 0.438) |
|  | Baseline SH ~ Follow-up 1 restricting | 0.000 (-0.169, 0.169) |
|  | Baseline SH ~ Follow-up 1 SI | 0.158 (-0.038, 0.355) - |
|  | Follow-up 1 SH ~ Follow-up 2 restricting | 0.020 (-0.186, 0.147) |
|  | Follow-up 1 SH ~ Follow-up 2 SI | 0.189 (-0.036, 0.413) - |
|  | Baseline SI ~ Follow-up 1 restricting | 0.034 (-0.194, 0.126) |
|  | Baseline SI ~ Follow-up 1 SH | 0.014 (-0.161, 0.190) - |
|  | Follow-up 1 SI ~ Follow-up 2 restricting | 0.066 (-0.227, 0.096) |
|  | Follow-up 1 SI ~ Follow-up 2 SH | 0.068 (-0.143, 0.280) |
| Individuals diagnosed with an eating disorder | Baseline purging ~ Follow-up 1 purging | 0.632 (0.500, 0.763) |
|  | Follow-up 1 purging ~ Follow-up 2 purging | 0.654 (0.531, 0.776) |
|  | Baseline SH ~ Follow-up 1 SH | 0.583 (0.426, 0.740) |
|  | Follow-up 1 SH ~ Follow-up 2 SH | 0.556 (0.376, 0.737) |
|  | Baseline SI ~ Follow-up 1 SI | 0.729 (0.602, 0.856) |
|  | Follow-up 1 SI ~ Follow-up 2 SI | 0.682 (0.529, 0.836) - |
|  | Baseline purging ~ Follow-up 1 SH | 0.016 (-0.204, 0.172) |
|  | Baseline purging ~ Follow-up 1 SI | 0.001 (-0.186, 0.188) |
|  | Follow-up 1 purging ~ Follow-up 2 SH | 0.073 (-0.131, 0.277) - |
|  | Follow-up 1 purging ~ Follow-up 2 SI | 0.026 (-0.236, 0.183) |
|  | Baseline SH ~ Follow-up 1 purging | 0.079 (-0.091, 0.249) |
|  | Baseline SH ~ Follow-up 1 SI | 0.141 (-0.054, 0.336) |
|  | Follow-up 1 SH ~ Follow-up 2 purging | 0.049 (-0.120, 0.219) |
|  | Follow-up 1 SH ~ Follow-up 2 SI | 0.170 (-0.055, 0.394) |
|  | Baseline SI ~ Follow-up 1 purging | 0.014 (-0.141, 0.169) |
|  | Baseline SI ~ Follow-up 1 SH | 0.018 (-0.163, 0.198) |
|  | Follow-up 1 SI ~ Follow-up 2 purging | 0.092 (-0.079, 0.263) |
|  | Follow-up 1 SI ~ Follow-up 2 SH | 0.051 (-0.166, 0.268) |
| Individuals diagnosed with an eating disorder | Baseline bingeing ~ Follow-up 1 bingeing | 0.745 (0.632, 0.859) |
|  | Follow-up 1 bingeing ~ Follow-up 2 bingeing | 0.663 (0.548, 0.777) |
|  | Baseline SH ~ Follow-up 1 SH | 0.573 (0.415, 0.732) |
|  | Follow-up 1 SH ~ Follow-up 2 SH | 0.561 (0.383, 0.740) |
|  | Baseline SI ~ Follow-up 1 SI | 0.731 (0.604, 0.858) |
|  | Follow-up 1 SI ~ Follow-up 2 SI | 0.673 (0.511, 0.835) |
|  | Baseline bingeing ~ Follow-up 1 SH | 0.088 (-0.075, 0.251) - |
|  | Baseline bingeing ~ Follow-up 1 SI | 0.069 (-0.233, 0.096) - |
|  | Follow-up 1 bingeing ~ Follow-up 2 SH | 0.126 (-0.301, 0.050) |
|  | Follow-up 1 bingeing ~ Follow-up 2 SI | 0.107 (-0.073, 0.286) - |
|  | Baseline SH ~ Follow-up 1 bingeing | 0.089 (-0.255, 0.078) |
|  | Baseline SH ~ Follow-up 1 SI | 0.152 (-0.042, 0.346) - |
|  | Follow-up 1 SH ~ Follow-up 2 bingeing | 0.102 (-0.274, 0.071) |
|  | Follow-up 1 SH ~ Follow-up 2 SI | 0.162 (-0.065, 0.389) - |
|  | Baseline SI ~ Follow-up 1 bingeing | 0.086 (-0.245, 0.074) - |
|  | Baseline SI ~ Follow-up 1 SH | 0.008 (-0.186, 0.170) - |
|  | Follow-up 1 SI ~ Follow-up 2 bingeing | 0.106 (-0.288, 0.076) |
|  | Follow-up 1 SI ~ Follow-up 2 SH | 0.080 (-0.139, 0.299) |

*Note*. CIs= confidence intervals; EDs= eating disorders; SH= self-harm; SI= suicidal ideation.

Supplementary Table 5. Results of the mediation analyses.

| Predictor | Outcome | Mediator | Indirect effect | SE | Z-value | P-value | Total |
| --- | --- | --- | --- | --- | --- | --- | --- |
| TEDS | | | | | | | |
| Age 16 EDs | Age 26 SI | Love and relationships | 0.005 | 0.005 | 0.888 | 0.375 | 0.093 |
| Age 16 EDs | Age 26 SI | Community | 0.007 | 0.004 | 1.605 | 0.109 | 0.083 |
| Age 16 EDs | Age 26 SI | Relationship with twin | 0.007 | 0.004 | 1.907 | 0.056 | 0.099 |
| Age 16 EDs | Age 26 SI | Count of life events | 0.023 | 0.007 | 3.477 | 0.001 | 0.089 |
| COPING | | | | | | | |
| Total EDs | | | | | | | |
| Baseline EDs | Follow-up 2 SI | Loneliness | 0.015 | 0.030 | 0.500 | 0.617 | 0.251 |
| Baseline EDs | Follow-up 2 SI | Relationship loss-related grief | 0.002 | 0.100 | 0.238 | 0.812 | 0.248 |
| Restricting | | | | | | | |
| Baseline restricting | Follow-up 2 SI | Loneliness | 0.151 | 0.091 | 1.666 | 0.096 | -0.065 |
| Baseline restricting | Follow-up 2 SI | Relationship loss-related grief | 0.020 | 0.032 | 0.633 | 0.537 | -0.110 |
| Baseline restricting | Follow-up 2 SI | Positive health behaviours | 0.014 | 0.028 | 0.512 | 0.609 | -0.065 |
| Purging | | | | | | | |
| Baseline purging | Follow-up 2 SI | Loneliness | -0.093 | 0.079 | -1.169 | 0.242 | -0.028 |
| Baseline purging | Follow-up 2 SI | Relationship loss-related grief | -0.034 | 0.044 | -0.774 | 0.439 | -0.025 |
| Baseline purging | Follow-up 2 SI | Positive health behaviours | -0.008 | 0.024 | -0.344 | 0.731 | -0.028 |
| Baseline purging | Follow-up 2 SI | Negative health behaviours | 0.008 | 0.030 | 0.272 | 0.785 | -0.028 |
| Bingeing | | | | | | | |
| Baseline bingeing | Follow-up 2 SI | Loneliness | 0.112 | 0.085 | 1.318 | 0.188 | 0.016 |
| Baseline bingeing | Follow-up 2 SI | Relationship loss-related grief | 0.023 | 0.035 | 0.674 | 0.500 | -0.026 |
| Baseline bingeing | Follow-up 2 SI | Positive health behaviours | 0.017 | 0.300 | 0.565 | 0.572 | 0.016 |
| Baseline bingeing | Follow-up 2 SI | Negative health behaviours | 0.006 | 0.024 | 0.232 | 0.816 | 0.016 |

*Note*. EDs= eating disorders; SI= suicidal ideation; SE= standard error.

Supplementary Figures

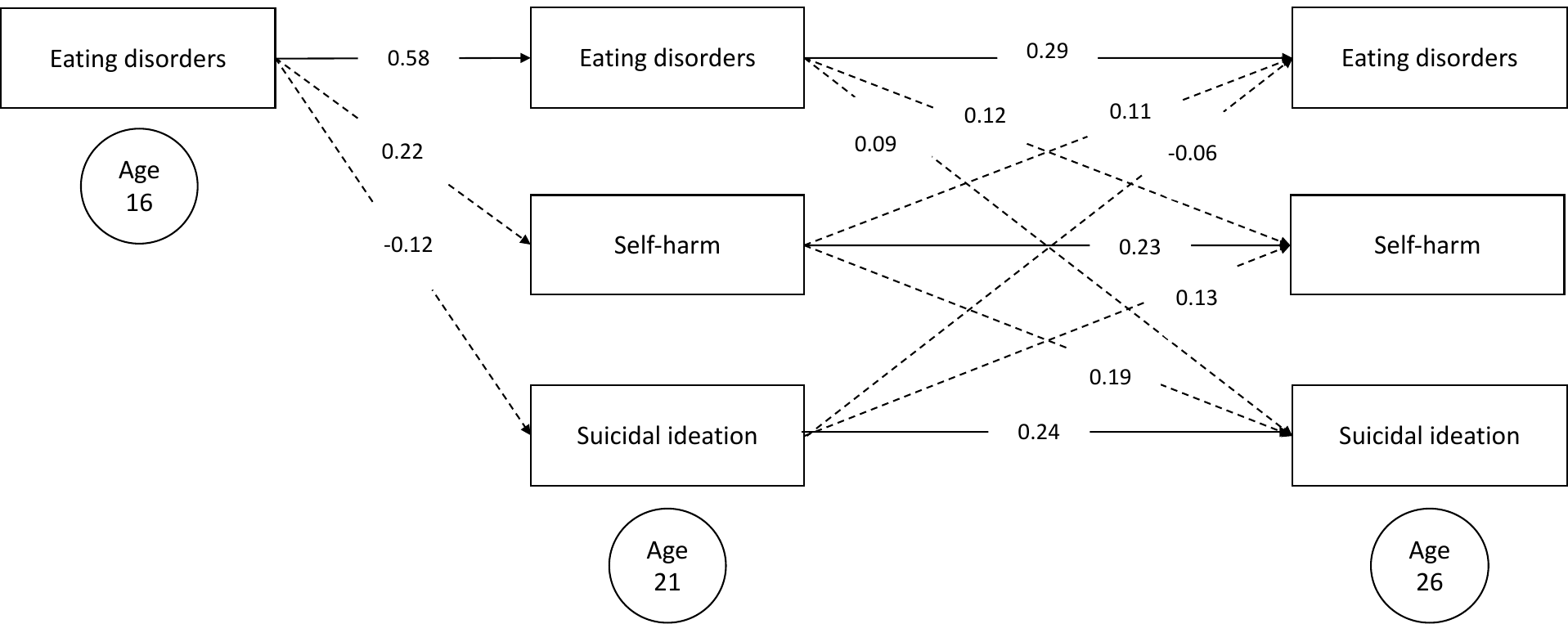

Supplementary Figure 1. The longitudinal path diagram of the individuals in the TEDS sample who have been diagnosed with any eating disorder (N=195). The models present the autoregressive longitudinal paths between symptoms of eating disorders, self-harm and suicidal ideation, as well as cross-trait longitudinal paths between the variables.

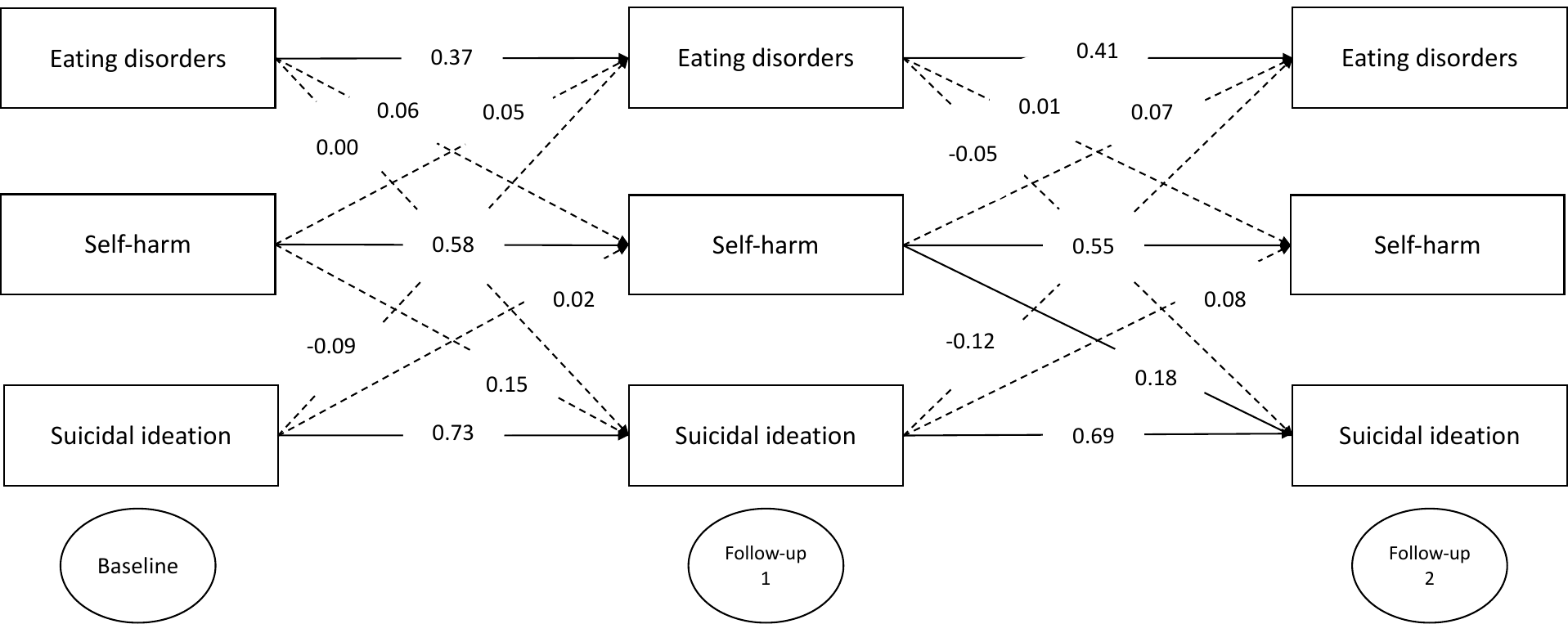

Supplementary Figure 2. The longitudinal path diagram of the individuals in the TEDS sample who have been diagnosed with any eating disorder (N=87). The models present the autoregressive longitudinal paths between symptoms of eating disorders, self-harm and suicidal ideation, as well as cross-trait longitudinal paths between the variables.

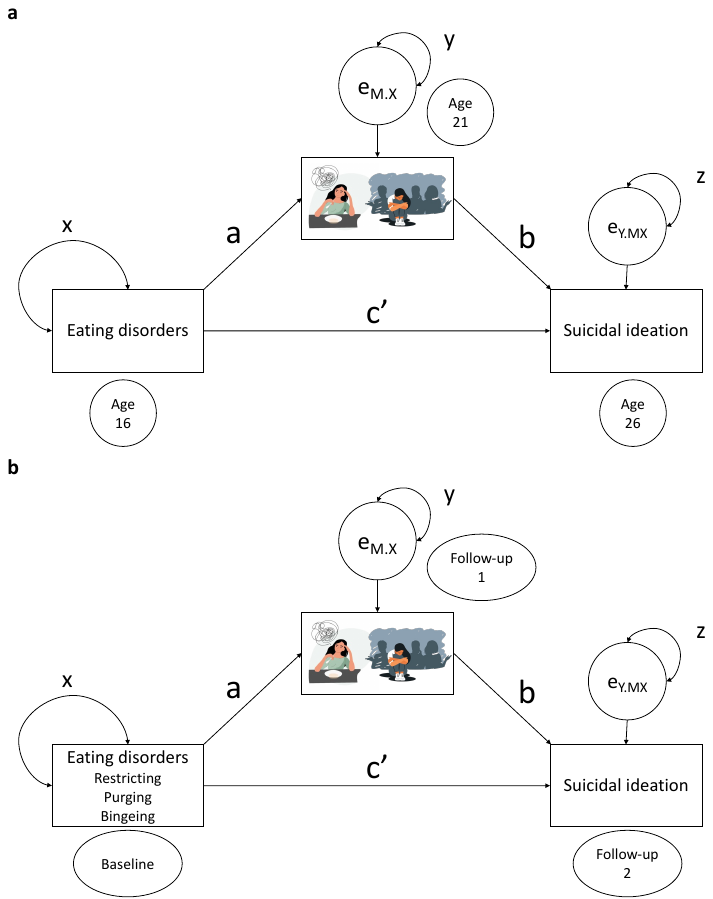

Supplementary Figure 3. Mediation diagrams. The model presents the mediation of symptoms of eating disorders on suicidal ideation, mediated by the psychosocial factors in TEDS (**a**) and multi-group models in COPING (**b**) samples. Circles indicate residuals. Parameters a, b and c represent regression weights. Parameters x, y and z represent variance parameters.

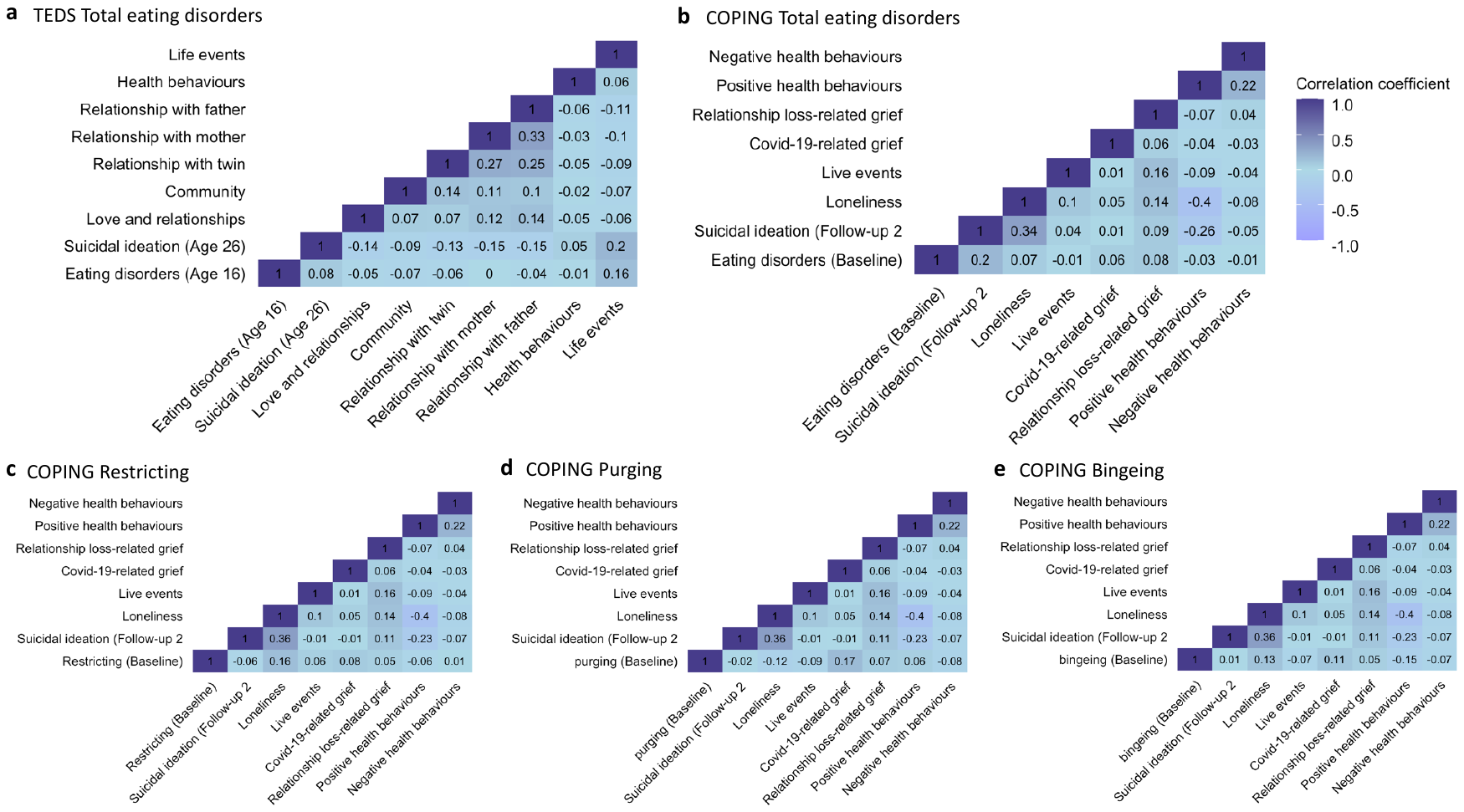

Supplementary Figure 4. Correlations between symptoms of eating disorders at the first timepoint and suicidal ideation at the final timepoint in TEDS (**a**) and COPING (**b**-**e**) samples.
